# In Vivo K-edge Imaging on a Clinical Dual-source PCCT: Feasibility of Gadolinium Applications in a Porcine Model

**DOI:** 10.64898/2026.09.07.26362472

**Authors:** Martin V. Rybertt, Leening P. Liu, Pooyan Sahbaee, George McClung, Manoj Mathew, Peter B. Noël, Ali H. Dhanaliwala

## Abstract

**Objectives:** To evaluate the feasibility of in vivo gadolinium K-edge imaging on a clinical dual-source photon-counting CT (PCCT) system in a porcine model and to assess its performance for dynamic contrast-enhanced imaging, functional biliary imaging, and dual-contrast (bi-phasic) imaging.

**Materials and Methods:** A single healthy Yorkshire-Landrace pig was imaged on a clinical dual-source PCCT system (140 kVp; energy thresholds 20/55/72/90 keV; CTDI_vol_ 12 mGy). Gadolinium K-edge material-specific maps were generated with a calibration-based least-squares material decomposition. Three imaging protocols were evaluated for feasibility: (A) contrast dose testing for dynamic imaging with a hepatobiliary-clearing gadolinium contrast agent (0.2 and 0.8 mL/kg); (B) functional biliary imaging with dynamic and delayed acquisitions; and (C) dual-contrast imaging with simultaneous iodine and gadolinium decomposition. Attenuation (HU) and material concentrations (mg/mL) were measured from regions of interest.

**Results:** Gadolinium was quantifiable in material-specific maps at a contrast dose of 0.8 mL/kg (arterial aortic signal, 114 ± 4 HU and 1.4 ± 0.3 mg/mL; delayed bile duct, 111 ± 4 HU and 4.3 ± 0.4 mg/mL). Hepatobiliary excretion was tracked over time, with bile duct gadolinium plateauing near 3.3 mg/mL approximately 15 minutes after injection. In the dual-contrast acquisition, arterial gadolinium (2.5 ± 0.7 mg/mL) and portal venous iodine (1.9 ± 0.7 mg/mL) were separated within the liver, and both agents were separated within the ureters.

**Conclusions:** In this proof of concept, gadolinium K-edge imaging was feasible on a clinical dual-source PCCT system for dynamic, functional biliary, and dual-contrast applications, supporting further development with improved material-decomposition sensitivity and confirmation in larger studies.

**Key Points:**

- Gadolinium K-edge imaging is feasible in vivo on a clinical dual-source photon-counting CT system, producing gadolinium-specific quantitative maps.
- Functional biliary imaging with gadoxetate disodium tracked hepatobiliary excretion over time, with biliary gadolinium plateauing approximately 15 minutes after injection.
- A dual-contrast acquisition separated gadolinium (arterial phase) from iodine (portal venous phase) within the liver and ureters, enabling simultaneous multi-phase, multi-agent imaging.

## Introduction

CT remains the workhorse imaging modality due to its rapid acquisition, relatively low dose, and high availability.^1^ As a cross-sectional modality, it is also an excellent screening tool in the emergency setting, allowing multiple organs to be assessed for abnormalities. Its versatility enables a wide range of examinations, including angiographic, dynamic, and multi-phasic studies. Clinical CT relies almost exclusively on iodinated contrast agents, showing a well-documented diagnostic utility.^2^ However, iodinated agents are contraindicated in patients with renal insufficiency, can cause allergic reactions, and provides limited enhancement of non-vascular structures like the biliary tree.^3^ Gadolinium-based agents—predominantly used in MRI—have been evaluated as an alternative CT contrast agent, but their weaker enhancement on conventional CT has limited their clinical utility for contrast-enhanced CT.^4–8^

In 2021, the first clinical photon-counting CT (PCCT) system was introduced,^9^ improving spatial resolution, reducing radiation dose, and enabling multi-bin imaging.^10,11^ Multi-bin imaging enables K-edge imaging, an advanced spectral technique that can uniquely characterize and quantify materials based on their K-edge energies. Named after the K-edge effect—a sharp increase in the attenuation of a material at a unique energy—this technique provides material-specific images with improved contrast and selective localization of contrast agents.^12–14^ Materials imaged with K-edge imaging must have a K-edge energy within the diagnostic CT energy range (40–90 KeV) and have a sufficient and similar number of photons at energies above and below its K-edge. Current FDA-approved iodinated contrast agents have a K-edge energy that is outside this range (33.2 keV). As a result, iodinated contrast agents are unsuitable for K-edge imaging in CT. Conversely, gadolinium-based contrast agents have a K-edge energy (50.2 keV) within the diagnostic CT energy range. Current dual-energy (DECT) spectral systems cannot perform K-edge imaging without assumptions as this technique requires three or more spectral channels to accurately describe the K-edge effect. Adopting contrast materials with suitable K-edge energies—such as gadolinium—as an alternative to iodine could broaden the clinical applications of contrast-enhanced CT. However, the feasibility and diagnostic utility of gadolinium K-edge imaging have yet to be established on a clinical PCCT system.

K-edge imaging has been extensively studied on pre-clinical PCCT systems, demonstrating several potential clinical applications. Early studies established the feasibility and benefits of dual-contrast K-edge imaging. By separating two distinct contrast agents within a single acquisition, dual-contrast imaging can reduce both radiation dose and inter-phase misregistration caused by patient motion.^15–17^ These advantages have been demonstrated for bi-phasic liver imaging,^18^ improved polyp visualization in colonography,^19^ and detection of thoracic endoleaks post-aortic repair.^20^ Material decomposition through K-edge imaging also improves differentiation of calcifications from neighboring CT enhancement—a known issue between iodine and calcium given their similar energy-dependent attenuation, improving diagnostic performance.^21^ Finally, K-edge imaging has the potential to enable CT-based molecular imaging. Current literature supports the use of nanoparticle-based contrast agents with K-edge materials, these can present lower toxicities, are highly customizable, and have improved targeting.^22–24^ Given this early success, it is essential to demonstrate its *in vivo* feasibility and evaluate the relevant imaging parameters before in-human evaluation.

Here, we evaluated the feasibility of *in vivo* gadolinium K-edge imaging on a clinical PCCT system in a porcine model. We assessed gadolinium-specific quantitative imaging across three complementary settings: dynamic contrast-enhanced imaging, functional biliary imaging, and dual-contrast (bi-phasic) imaging. Demonstrating *in vivo* K-edge imaging in a clinical PCCT would further expand its capabilities and facilitate its translation into routine clinical practice.

## Methods

### Experimental setup

For multi-bin imaging, we utilized a clinical PCCT system (NAEOTOM Alpha.Peak, Siemens Healthineers, Forchheim, Germany). This PCCT system can separate incident photons into four energy bins, allowing the reconstruction of up to four threshold images. In this study, for all gadolinium samples and injections we used gadoxetate disodium (0.25 mmol/mL, Eovist, Bayer Healthcare, Whippany, NJ), a gadolinium-based contrast agent that is excreted through the renal and biliary systems. Additionally, iopamidol (Isovue-370, Bracco Diagnostics, Milan, Italy), an iodinated contrast agent, was used for preparation of iodine stock solutions and for contrast injection during the dual-contrast protocol.

### Material decomposition

To isolate these agents, we employed a material decomposition approach described by Neumann et al.^25^ to perform *in vivo* K-edge imaging of gadolinium. This method leverages a set of expected values from a calibration to perform a least squares estimation that generates material-specific maps of K-edge contrast agents. During the calibration process, sample materials of a known concentration were prepared and scanned within a vendor-provided phantom that has a solid water background. Samples of water, iodine (13.5 mg/mL), and gadolinium (10.0 mg/mL) were scanned separately at varying object diameters (10–40 cm), tube currents (25–800 mA), and distances to isocenter (0–5 cm) to generate a set of expected values. During material decomposition, threshold input images were water corrected and served as inputs to the material decomposition via a least squares estimation to reconstruct material-specific images.

### Animal model

This animal study was approved by the local Institutional Animal Care and Use Committee (IACUC, ID: 807750). Over the course of the study, the same healthy Yorkshire-Landrace pig was imaged during two sessions at 3 and 6 months of age, corresponding to 20 and 40 kg of weight, respectively. At 3 months of age and 20 kg, the pig was subjected to dynamic contrast-enhanced imaging for a contrast dose evaluation (A) and dual-contrast imaging (C). At 6 months of age and 40 kg, the pig was subjected to a functional biliary imaging protocol (B). The pig was fasted prior to each imaging session. Sedation was initiated with an intramuscular injection of the following preanesthetic cocktail: midazolam (Versed, Hospira, Lake Forest, IL, USA, 0.2 mg/kg), dexmedetomidine (Dexdomitor, Zoetis, Parsippany, NJ, USA, 0.2 μg/kg), and butorphanol (Torbugesic, Zoetis, Parsippany, NJ, USA, 0.2 mg/kg). Before imaging, an auricular intravenous (IV) catheter was placed. Anesthesia was induced with an IV propofol (PropoFlow, Zoetis, Parsippany, NJ, 4 mg/kg titrated to effect) injection followed by 2–3% inhaled isoflurane (Isospire, Dechra Pharmaceuticals PLC, Northwich, United Kingdom) after intubation. The pig was then placed on the CT table in a prone position for subsequent imaging. Prior to contrast injection, a non-contrast acquisition was performed provide an anatomical reference and confirm the absence of contrast. Anesthesia was reversed after completion of the imaging session.

### Imaging protocols

#### A. Contrast dose evaluation

To evaluate the gadolinium contrast dose, the pig (20 kg) was injected with 0.2 mL/kg—twice the suggested contrast dose for clinical use—of the gadolinium-based contrast agent after the pre-contrast baseline scan. Contrast was injected at a rate of 3 mL/s followed by a saline flush of 10 mL using a dual-head power injector (MEDRAD Stellant Flex, Bayer LLC, Whippany, NJ, USA). After beginning the injection, images were acquired every 6.5 seconds for 24 acquisitions. Following a 30-minute washout period, the protocol was repeated with a 0.8 mL/kg contrast dose (Figure 1A).

**Figure 1.**
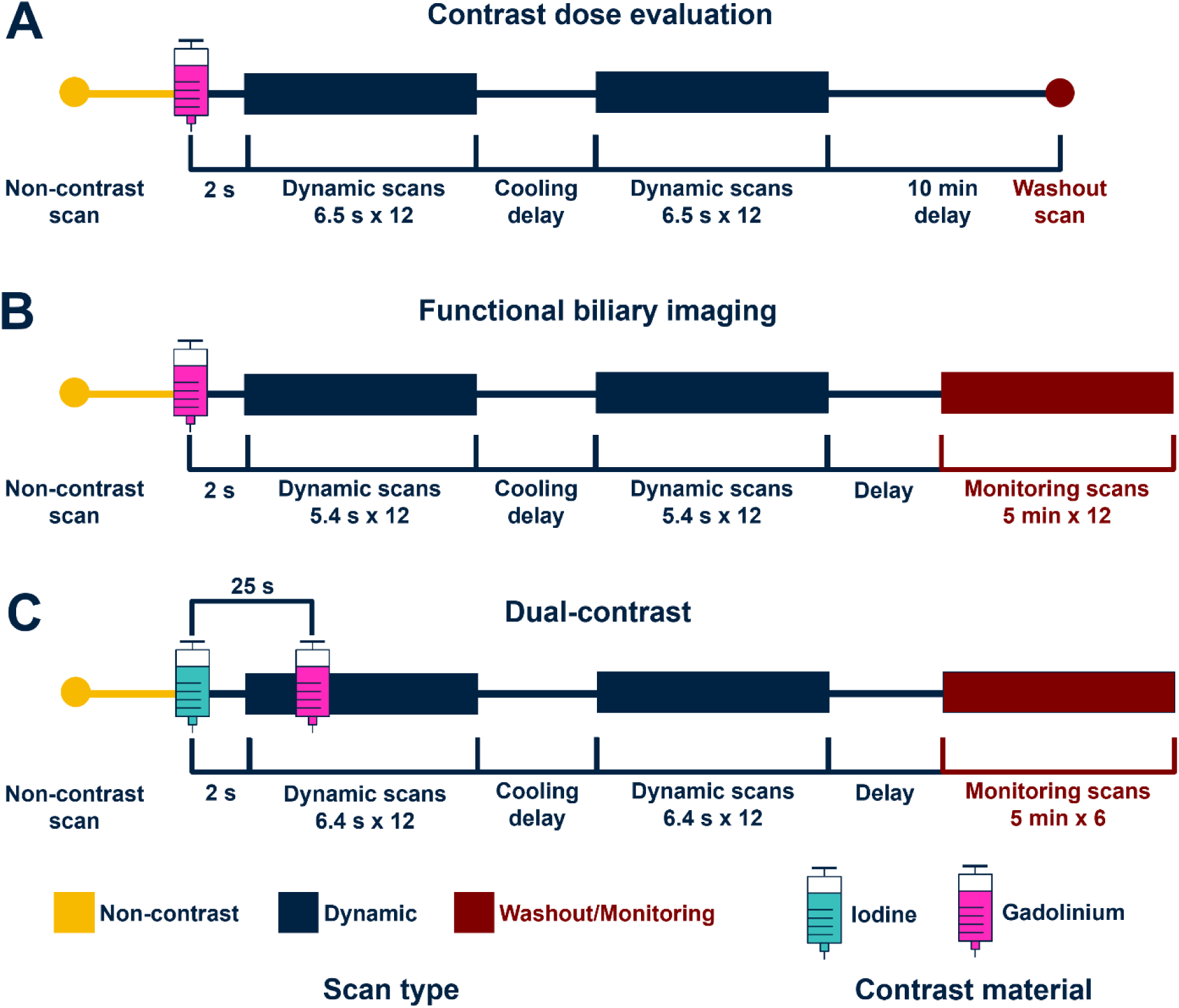
Contrast injection timing and acquisition protocols. (A) Dose evaluation. First, a 0.2 mL/kg gadolinium-based contrast injection was delivered, followed by 0.8 mL/kg dose after a 30-minute washout. (B) Biliary imaging. A 1 mL/kg gadolinium contrast was injected, with subsequent dynamic imaging. Then, scans were obtained every 5 minutes to monitor excretion of gadolinium onto the bile duct and gallbladder. (C) Dual-contrast. 0.75 mL/kg iodine-based contrast followed by 1 mL/kg gadolinium-based contrast after a 25 second delay, to capture both the portal venous phase (iodine) and the arterial phase (gadolinium) simultaneously.

#### B. Biliary imaging

To investigate biliary imaging, the pig (40 kg) first received a baseline abdominopelvic pre-contrast scan. Subsequently, a 1 mL/kg contrast dose was injected at 4 mL/s followed by 10 mL of saline. Scans were acquired every 5.4 seconds, starting 2 seconds after injection for 24 acquisitions and then every five minutes for an hour. The protocol was repeated after 48 hours—labeled day 1 and day 3, respectively—to assess the consistency of the excretion pattern (Figure 1B).

#### C. Dual-contrast imaging

To investigate multi-contrast imaging, the pig (20 kg) was first injected with a 20 mL iodinated contrast test bolus to determine the injection timing of the portal venous phase. For the protocol, iodinated contrast (0.75 mL/kg) was injected at a rate of 4 mL/s by a dual-head power injector. After a 25 second delay, gadolinium-based contrast (1 mL/kg) was injected at the same rate to provide arterial contrast. Tissue-mimicking rods of 10.0 mg/mL of iodine (Sun Nuclear, Melbourne, FL, USA) and water, as well as a 10.0 mg/mL dilution of gadolinium (Eovist) were placed alongside the abdomen as a control. Scans were performed every 6.4 seconds, starting 2 seconds after injection for a total of 24 repetitions (Figure 1C).

### Image acquisition and reconstruction

All scans were performed on a clinical PCCT system (NAEOTOM Alpha.Peak; Siemens Healthineers, Forchheim, Germany) with a tube voltage of 140 kVp, an energy bin configuration of 20/55/72/90 keV, and a volumetric CT dose index (CTDI_vol_) of 12 mGy. CT raw projection data was collected and loaded into a vendor-specific workstation (ReconCT, Siemens Healthineers, Forchheim, Germany) to generate conventional images and spectral post-processing (SPPs) files. Vendor-provided software (eXamine, Siemens Healthineers, Forchheim, Germany) was used to process SPP files and generate material-specific maps of water, iodine, and gadolinium. Images were reconstructed with a quantitative reconstruction kernel (Qr40), a denoising (QIR) level of 3, and a matrix size of 512 x 512 px (Table 1).

**Table 1.**
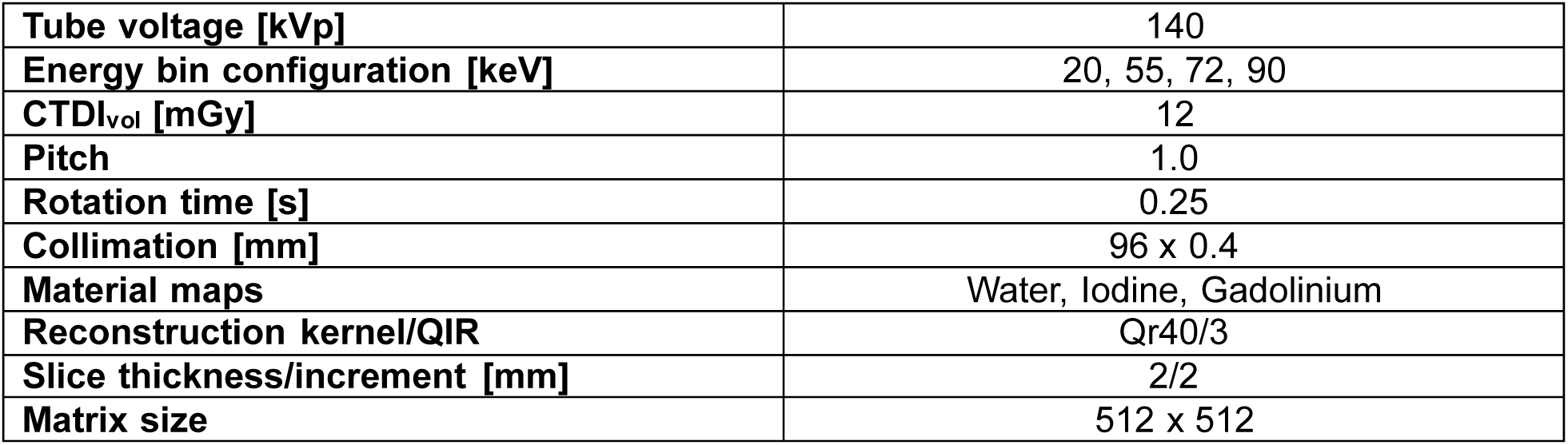
Acquisition and reconstruction parameters.

### Image analysis

Regions of interest (ROIs) were placed on conventional images at select anatomical locations. ROI locations were then utilized to measure the same coordinates on gadolinium maps. Mean and standard deviation of the ROIs were used to collect attenuation and concentration at each location for each timepoint. Using this information, the dynamic enhancement phases such as arterial and portal venous phases were identified by the timing of their peak enhancement. For the contrast dose evaluation (A), ROIs were placed on the aorta and liver parenchyma. For biliary imaging (B), ROIs were also placed on the gallbladder and bile ducts to measure excretion. In addition, excretory functional timepoints such as time-to-peak and time-to-plateau within the biliary tree were estimated. For the dual-contrast protocol (C), quantification error using the control samples was calculated as the difference between expected and measured mean concentration within the sample.

## Results

### A. Contrast dose

*In vivo* quantitative imaging of gadolinium was successful at larger contrast doses. On the reference non-contrast images, no quantifiable or identifiable gadolinium enhancement was observed. During dynamic imaging, IV injection of gadolinium produced enhancement of the aorta, bile duct, and bowel within 10 minutes, with the degree of enhancement varying by contrast dose. The 0.2 mL/kg injection presented discernible contrast during the arterial phase at the aorta only in conventional images where attenuation measured 60 ± 6 HU. Bile duct and bowel excretion were both detected in conventional images 10 minutes post-injection, but only bowel excretion could be differentiated in the gadolinium map (Figure 2). In comparison, when using a 0.8 mL/kg contrast dose, arterial aortic enhancement was visible and measured 114 ± 4 HU and 1.4 ± 0.3 mg/mL in conventional and gadolinium maps, respectively. Furthermore, residual gadolinium excretion in the bowel from the previous injection was observable. The delayed phase showed increased enhancement in the bile duct compared to the 0.2 mL/kg injection, reaching a gadolinium signal of 111 ± 4 HU and concentration of 4.3 ± 0.4 mg/mL for conventional and gadolinium images, respectively.

**Figure 2.**
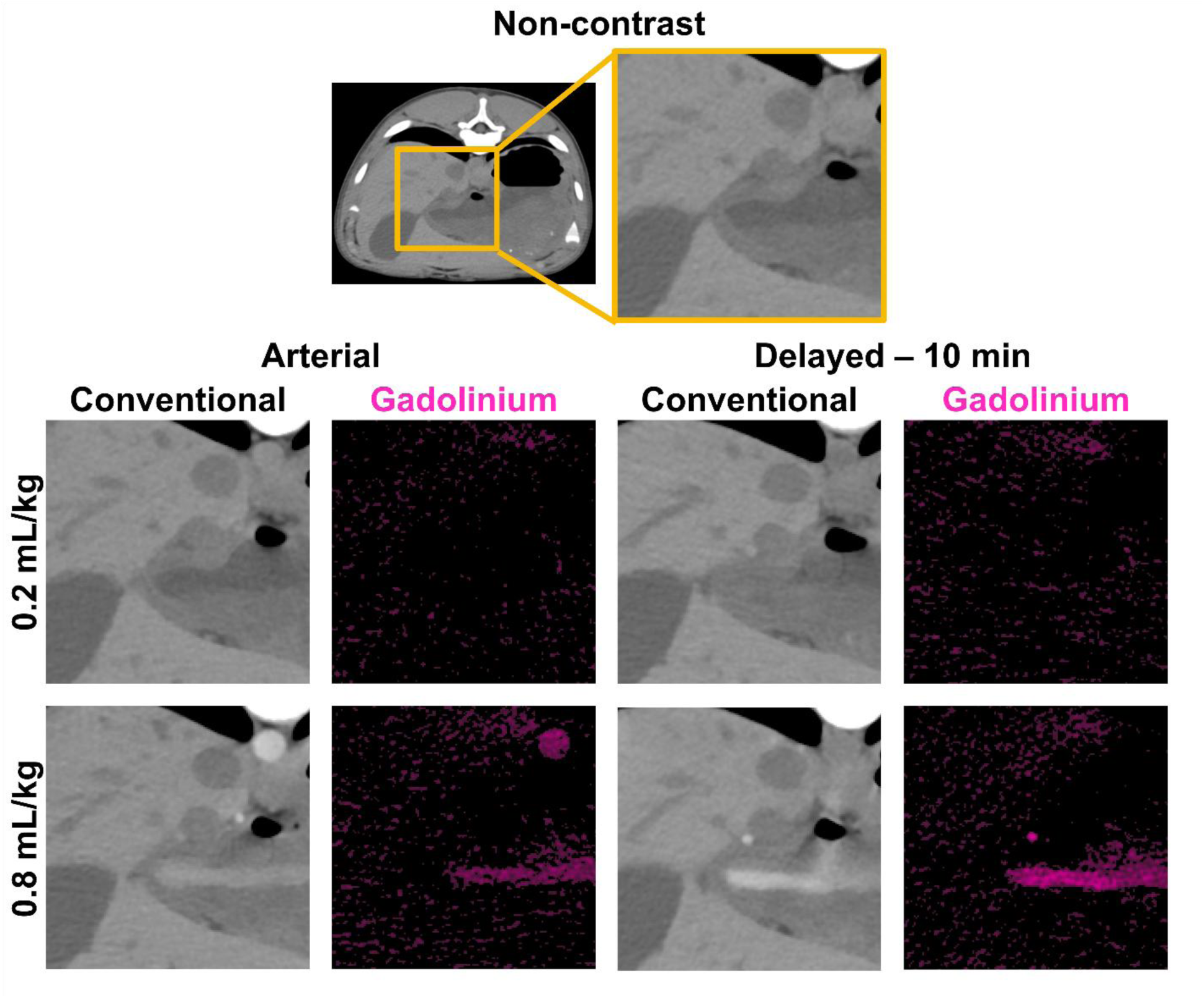
Evaluation of feasibility and contrast dose. Non-contrast axial images were used for comparison with post-injection acquisitions. A 0.2 mL/kg injection of gadolinium-based contrast did not demonstrate vascular enhancement during its arterial phase for conventional (WL/WW: 40/300 HU) and gadolinium-specific (WL/WW: 1.5/3 mg/mL) images. Whereas the delayed phase (10 minutes post-contrast) presented bile duct enhancement on conventional images. The arterial phase of the 0.8 mL/kg injection of gadolinium resulted in contrast signal in both conventional and gadolinium images. Residual gadolinium contrast from the previous injection can be observed in the bile duct and bowel. After the delay, the gadolinium excretory pathway into the bile duct and bowel was visualized.

### B. Biliary imaging

Both dynamic and functional stages of the hepatobiliary clearance of gadolinium were identified and quantified using PCCT. On the baseline non-contrast images demonstrated no measurable gadolinium signal in the aorta, liver parenchyma, gallbladder, or bile ducts (Figure 3A). On the baseline non-contrast images demonstrated no measurable gadolinium signal in the aorta, liver parenchyma, gallbladder, or bile ducts (Figure 3A). Compared to the contrast dose experiment, the timing of the arterial phase was recorded at 13.2 seconds, likely owing to the larger pig weight. Aortic enhancement peaked at 110 ± 6 HU with a corresponding gadolinium concentration of 2.8 ± 0.9 mg/mL (Figure 3B) for the day 1 experiment. A distinct portal venous phase was not readily discernible under these scanning conditions, as the liver parenchyma showed a contrast increase lower than 15 HU or 0.5 mg/mL with respect to our baseline.

**Figure 3.**
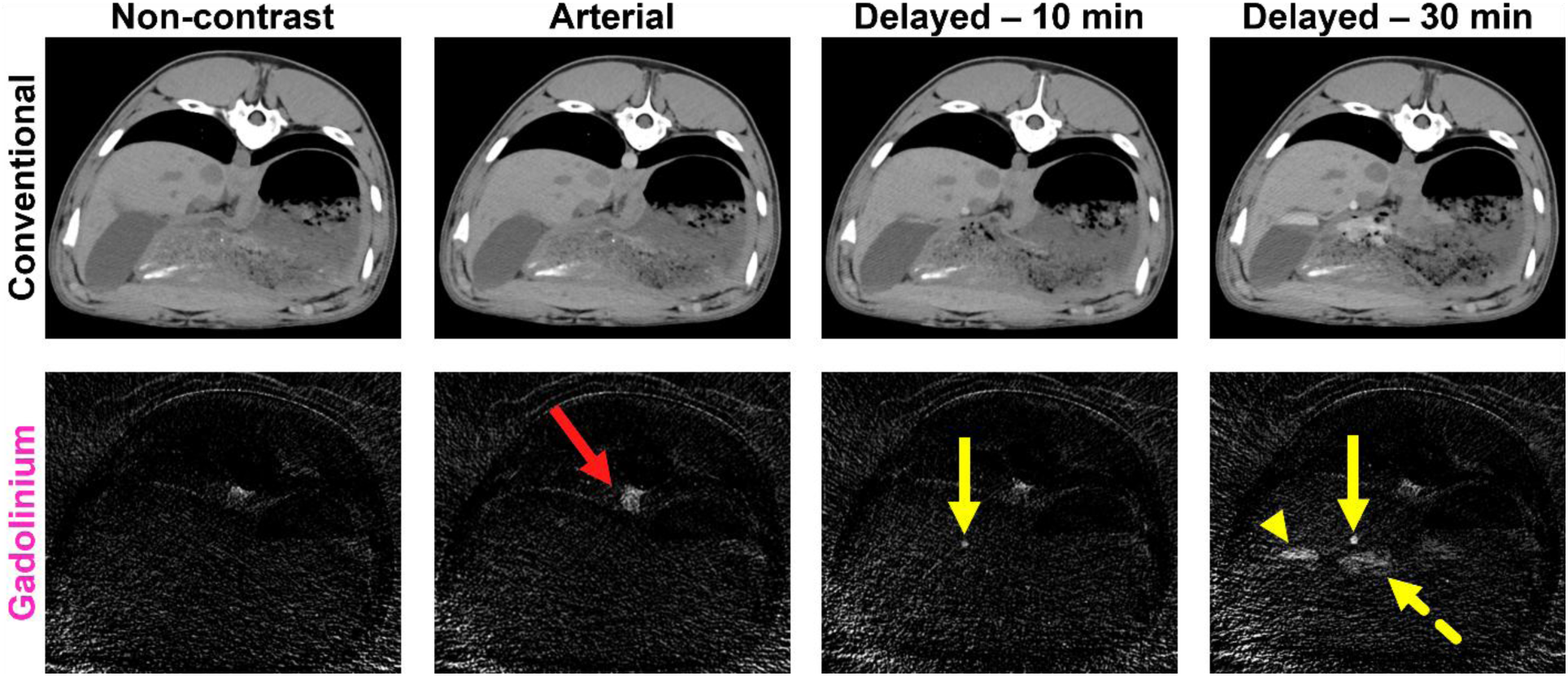
Biliary excretion of gadolinium. Non-contrast conventional (top) images displayed no gadolinium enhancement, corroborated by the lack of signal on gadolinium maps (bottom). Gadolinium contrast was first measured at the aorta during the arterial phase (red arrow), while the clearance of gadolinium into the bile duct (solid yellow arrow), followed by excretion into the gallbladder (yellow arrowhead) and bowel (dashed yellow arrow) became discernible after a delay. Conventional: WL/WW: 40/300 HU; Gadolinium: 3/6 mg/mL

Hepatobiliary excretion of gadolinium onto the biliary was consistent across experiment days. Gadolinium excretion to the bile duct became apparent 10 minutes post-injection (Figure 3C), measuring 3.3 ± 0.8 and 2.7 ± 1.0 mg/mL for day 1 and day 3 experiments, respectively. In parallel, concentration at the gallbladder was 0.7 ± 1.0 and 2.2 ± 0.8 mg/mL for each experiment, respectively. At 15 minutes post-injection, excretion of gadolinium onto the biliary system plateaued, stabilizing at 3.3 ± 0.9 and 3.0 ± 1.1 mg/mL in the bile ducts for experiment on day 1 and day 3 (Figure 4), respectively. In addition, the concentration of gadolinium within the gallbladder measured 3.2 ± 0.9 and 2.5 ± 0.9 mg/mL. After 30 minutes, the full excretory pathway for the gadolinium contrast agent was visualized, with simultaneous accumulation in the bile duct, gallbladder, and bowel (Figure 3D).

**Figure 4.**
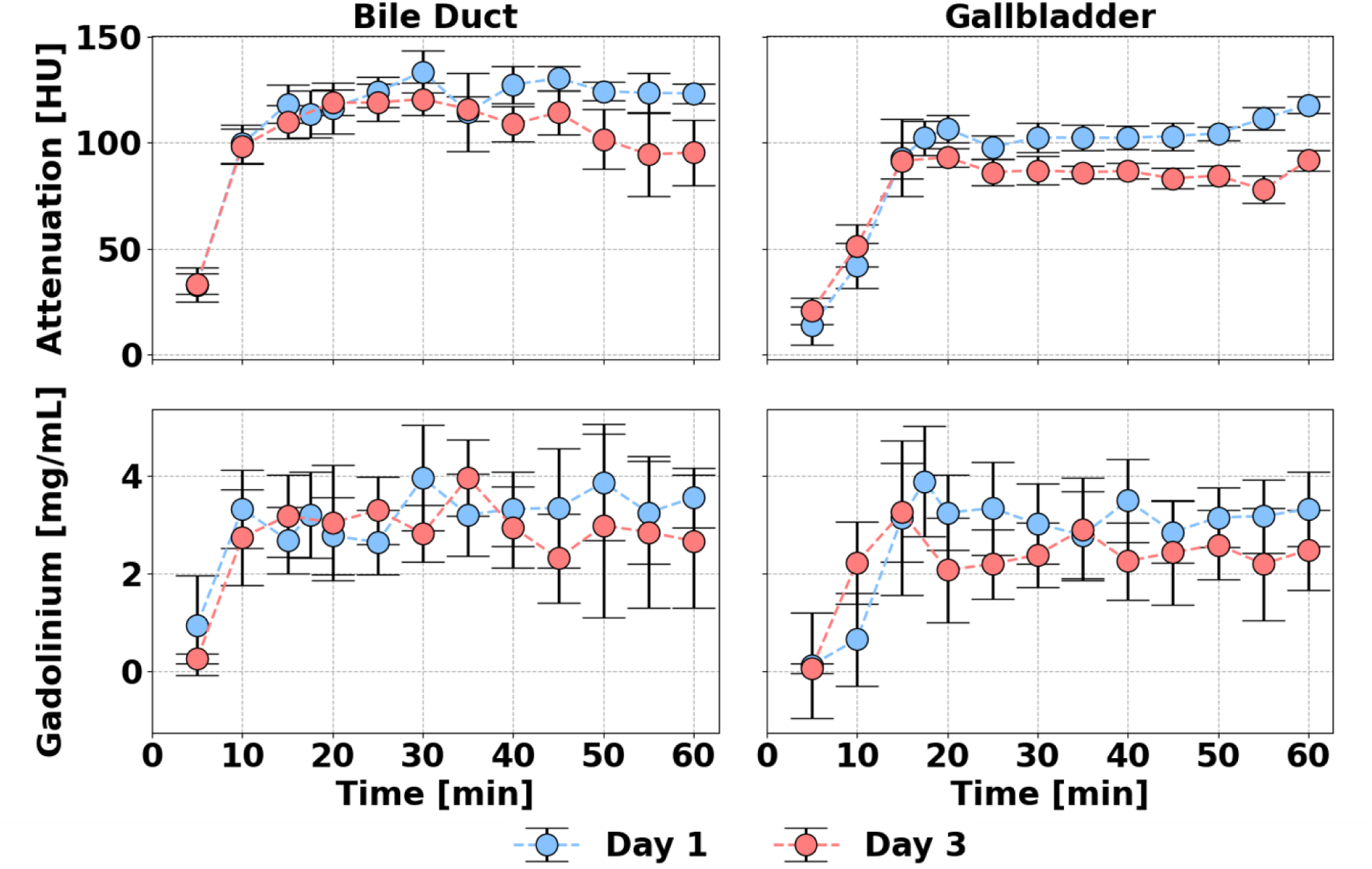
Enhancement of biliary structures over time on conventional and gadolinium images. Conventional and gadolinium images display an increase in contrast signal in the bile ducts and gallbladder ten minutes after the start of injection, with gadolinium concentration plateauing approximately 15 minutes after injection. Error bars describe the standard deviation (noise) of the region of interest.

**Figure 5.**
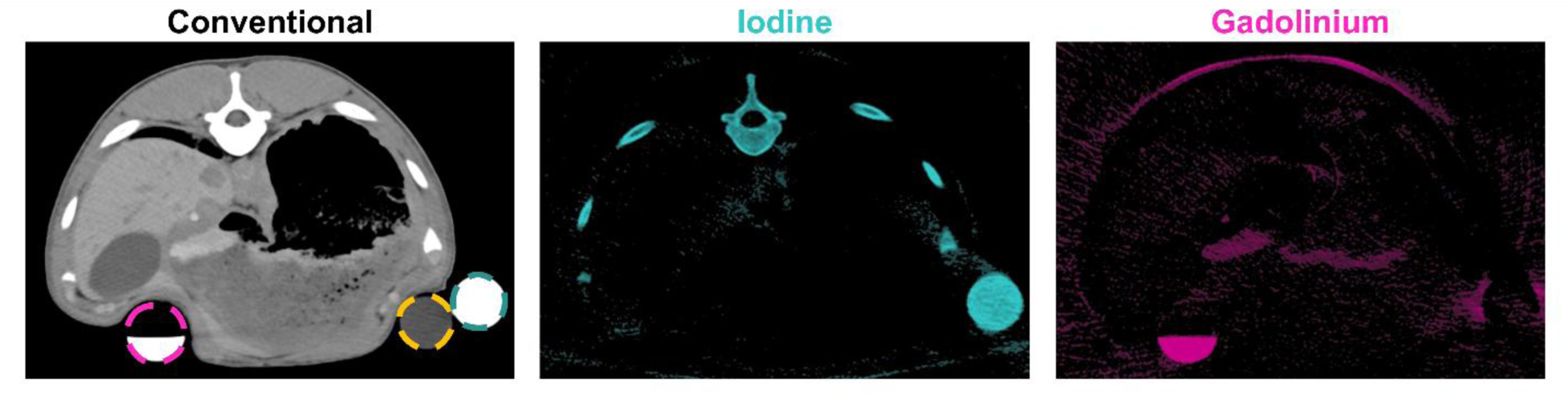
Dual-contrast imaging of iodine and gadolinium in a porcine model in separate volumes. Material decomposition results in correct localization of iodine (teal) and gadolinium (pink) in control material inserts and solutions. Whereas a solid water rod (gold) did not display enhancement on material-specific images. Conventional images (WL/WW: 40/300 HU) showed excretion of gadolinium into the bowel. This is confirmed by the lack of signal in iodine images (WL/WW: 6/12 mg/mL) and the high contrast in gadolinium images (WL/WW: 3/6 mg/mL).

### C. Dual-contrast imaging

The bi-phasic protocol displayed simultaneous visualization of the arterial (gadolinium) and portal venous (iodine) phases of the liver within a single acquisition. Conventional images were unable to separate iodine from gadolinium enhancement; however, iodine and gadolinium maps showed distinct localization of each contrast in the aorta and liver. From the same acquisition, arterial aortic enhancement of gadolinium reached 2.5 ± 0.7 mg/mL while iodine portal venous enhancement in the liver parenchyma was 1.9 ± 0.7 mg/mL.

Furthermore, *in vivo* material decomposition of iodine and gadolinium within the same volume was feasible. 30 minutes post-injection, iodine and gadolinium accumulated in the ureters and kidneys. Material-specific images presented clear separation of the two contrast agents within the ureters (Figure 6).

**Figure 6.**
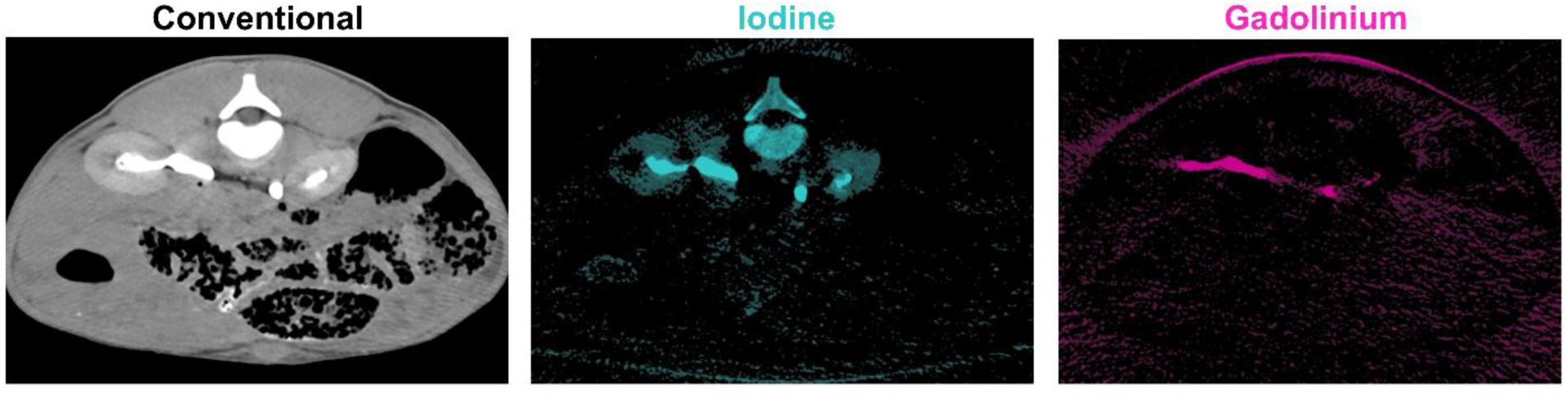
Simultaneous material decomposition of iodine and gadolinium within the ureters. Dual-contrast acquisition of iodine and gadolinium exhibited concurrent excretion of iodine and gadolinium through the kidneys. Iodine- and gadolinium-based contrast agents were not discernible from each other on conventional (WL/WW: 40/300 HU) images. Material-specific images of iodine and gadolinium indicate separation of iodine (WL/WW: 6/12 mg/mL) and gadolinium (WL/WW: 3/6 mg/mL) within the same volume.

## Discussion

In this study, we evaluated the feasibility of *in vivo* gadolinium K-edge imaging on a clinically available dual-source PCCT system in a porcine model. Across three imaging protocols, we found that (i) gadolinium can be quantified in material-specific maps at a dose of 0.8 mL/kg; (ii) hepatobiliary excretion of gadoxetate disodium could be tracked dynamically, with bile duct signal that is detectable and plateaus approximately 10 and 15 minutes after injection, respectively; and (iii) gadolinium and iodine could be separated within a single acquisition, both in the liver during their respective contrast phase and at the ureters during washout. These findings should be interpreted as an initial, single-animal proof-of-concept of *in vivo* K-edge imaging.

K-edge imaging has been demonstrated extensively on preclinical and research spectral PCTT systems, where it has enabled dual-contrast agent separation, molecular imaging of targeted nanoparticles, and functional material discrimination.^13,14,18,20,21,26,27^ The present work extends these capabilities to a clinically available dual-source PCCT platform using a calibration-based, prior-knowledge-free material decomposition.^25^ The performance of this approach was previously characterized in phantoms and a rat model.^28–30^ Thus, our study adds an early demonstration using both a clinical PCCT system and contrast agents in a large animal model for three candidate applications.

The contrast dose dependence of gadolinium detectability reflects the sensitivity limit of material decomposition at low K-edge material concentrations. The gadolinium-based contrast agent utilized in this study, gadoxetate disodium, carries a relatively low elemental gadolinium concentration (39.3 mg/mL) compared with the iodine concentration of iodinated agents (>300 mg/mL). Consequently, the *in vivo* gadolinium signal was quantitative by using a higher contrast dose of 0.8 mL/kg. However, when using the lower 0.2 mL/kg dose, the gadolinium signal likely fell below the noise floor of the decomposition. This is consistent with a human gadoxetate DECT study in which hepatic parenchymal enhancement was not observed.^31^ Because DECT is unable to describe the K-edge effect, our results from a K-edge sensitive system (PCCT) further emphasize the challenge of detecting gadolinium in the liver parenchyma using clinically permissible doses of the contrast agent in CT. Thus, to improve quantitative gadolinium K-edge imaging at the current dosage will accordingly require gains in decomposition sensitivity. This increase can be achieved by developing dedicated denoising, employing projection-based decomposition, or formulating higher concentration gadolinium agents. Furthermore, the energy threshold configuration used here (20/55/72/90 keV) does not place a threshold immediately adjacent to the K-edge of gadolinium (50.2 keV), which may further limit sensitivity at lower contrast doses. Although the effect of threshold placement was not evaluated in this study, an optimization of threshold energies could improve the decomposition sensitivity needed for clinical translation.

For functional biliary imaging, gadoxetate disodium—a hepatobiliary-excreted agent used routinely in MRI—permitted CT-based visualization of biliary clearance despite negligible parenchymal enhancement. The observed time course (detectability by ∼10 minutes, plateau by ∼15 minutes) indicates that dynamic, quantitative biliary CT is feasible and could complement MR cholangiography, particularly where MRI or iodinated contrast is contraindicated.^32^ Our measured gallbladder concentrations were lower than values predicted from a phantom model,^33^ plausibly because continuous excretion during dynamic imaging prevents full biliary accumulation. This explanation is provisional and was not directly tested. Furthermore, as these observations are derived from a single animal, the apparent stability of the excretion time course should be regarded as preliminary. Confirmation will require evaluation across multiple animals as well as employing models with biliary pathologies.

Dual-contrast imaging is among the most compelling applications of K-edge imaging. Separating two agents within a single acquisition can reduce cumulative radiation dose and remove the inter-phase misregistration inherent to sequential multi-phase protocols. In this study, we separated gadolinium and iodine both within the liver and the ureters. Quantitatively, however, the error in the gadolinium control solution was high and inconsistent with previous phantom studies.^28,29^ Potential explanations include error within sample preparation and the known sensitivity of the decomposition approach to off-centering and beam hardening. Because the source of this difference could not be discerned retrospectively, we elected not to report the quantitative values of the control solutions and will explore improvements for the latter effects in future studies. Furthermore, given the lower errors observed in other evaluations of this decomposition approach,^25,29^ we do not believe this is caused by a systemic bias in the imaging process. Ren et al. reported reduced quantification error and noise for iodine–gadolinium dual-contrast photon-counting-detector CT using a volume constraint and deep-learning denoising.^15^ Although our decomposition already models noise covariance,^25^ incorporating dedicated denoising, anti-correlated noise modeling,^34^ or spectral iterative reconstruction^35^ would be expected to improve dual-contrast quantitative accuracy.

This study has several limitations. First and most fundamentally, it comprises a single healthy animal; we therefore report no measures of reproducibility or statistical inference, and the reported standard deviation values represent within-region-of-interest dispersion from single acquisitions rather than between-subject uncertainty. A larger cohort, representing different anatomies and pathologies, is necessary for generalizability of our findings. Second, quantifiable gadolinium imaging with K-edge imaging required doses above the current dose recommended in the instructions for use. Applying methods that improve sensitivity—as described previously—could reduce the required contrast dose for quantification. Third, decomposition relied on proprietary, image-based vendor software,^25^ which is subject to beam-hardening artifacts and has restricted reproducibility.

Future studies will compare the performance of alternative material decomposition approaches. In summary, gadolinium K-edge imaging is feasible in vivo on a clinical dual-source PCCT system and supports dynamic, functional biliary, and dual-contrast applications in this single-animal proof of concept. K-edge imaging has vast clinical potential: broadening the range of available CT contrast agents, enabling quantitative multi-agent imaging, and reducing radiation dose and misregistration in multi-phasic studies. Clinical translation will entail improvements in material decomposition sensitivity, optimization of acquisition protocols of the gadolinium K-edge, and validating *in vivo* performance in adequately powered studies. These results represent an early and encouraging step toward that goal.

## Data Availability

All data produced in the present study are available upon reasonable request to the authors

## Acknowledgment

This work was partly supported by Siemens Healthineers and by the National Institutes of Health (NIH) (R01EB035908).

